# Evaluating Large Language Models in Echocardiography Reporting: Opportunities and Challenges

**DOI:** 10.1101/2024.01.18.24301503

**Authors:** Chieh-Ju Chao, Imon Banerjee, Reza Arsanjani, Chadi Ayoub, Andrew Tseng, Jean-Benoit Delbrouck, Garvan C. Kane, Francisco Lopez-Jimenez, Zachi Attia, Jae K Oh, Bradley Erickson, Li Fei-Fei, Ehsan Adeli, Curtis Langlotz

**Affiliations:** Department of Cardiovascular Medicine, Mayo Clinic, Rochester, Minnesota; Stanford Institute for Human-Centered Artificial Intelligence, Palo Alto, California; Department of Radiology, Mayo Clinic, Scottsdale, Arizona; Department of Cardiovascular Diseases, Mayo Clinic Arizona, Scottsdale, Arizona; Department of Cardiovascular Medicine, Mayo Clinic Florida, Jacksonville, Florida; Center for Artificial Intelligence in Medicine and Imaging (AIMI), Stanford University, Palo Alto, California; Department of Radiology, Mayo Clinic, Rochester Minnesota

**Keywords:** Artificial Intelligence, Large Language Model, Echocardiography, Quality, Cardiovascular Imaging

## Abstract

**Background:** The increasing need for diagnostic echocardiography (echo) tests presents challenges in preserving the quality and promptness of reports. While Large Language Models (LLMs) have proven effective in summarizing clinical texts, their application in echo remains underexplored.

**Aims:** To evaluate open-source LLMs in echo report summarization.

**Methods:** Adult echo studies conducted at the Mayo Clinic from January 1, 2017, to December 31, 2017, were categorized into two groups: development (all Mayo locations except Arizona) and Arizona validation sets. We adapted open-source LLMs (Llama-2, MedAlpaca, Zephyr, and Flan-T5) using In-Context Learning (ICL) and Quantized Low-Rank Adaptation (QLoRA) fine-tuning for echo report summarization from “Findings” to “Impressions.” Against cardiologist-generated Impressions, the models’ performance was assessed both quantitatively with automatic metrics and qualitatively by cardiologists.

**Results:** The development dataset included 97,506 reports from 71,717 unique patients, predominantly male (55.4%), with an average age of 64.3±15.8 years. EchoGPT, a QLoRA fine-tuned Llama-2 model, outperformed other LLMs with win rates ranging from 87% to 99% in various automatic metrics, and produced reports comparable to cardiologists in qualitative review (significantly preferred in conciseness (p< 0.001), with no significant preference in completeness, correctness, and clinical utility). Correlations between automatic and human metrics were fair to modest, with the best being RadGraph F1 scores versus clinical utility (r=0.42) and automatic metrics showed insensitivity (0-5% drop) to changes in measurement numbers.

**Conclusions:** EchoGPT can generate draft reports for human review and approval, helping to streamline the workflow. However, scalable evaluation approaches dedicated to echo reports remains necessary.

**Clinical Perspectives:** 1. What is new?

- This study evaluated multiple open-source LLMs and different model adaptation methods in echocardiography report summarization.
- The resulting system, EchoGPT, can generate echo reports comparable in quality to cardiologists.
- Future metrics for echo report quality should emphasize factual correctness, especially on numerical measurements.

2. What are the clinical implications?

- EchoGPT system demonstrated the potential of introducing LLMs into echocardiography practice to generate draft reports for human review and approval.

## Introduction

Echocardiography (echo) is the mainstay imaging modality in the current practice of cardiology(1), providing vital, non-invasive assessments of heart anatomy and physiology to guide clinical decisions(2). In the past decade, the rising demand for diagnostic echo tests(3) has posed significant challenges in maintaining the quality and timeliness of diagnostic reports(4–7), underscoring the necessity for automated solutions to enhance both efficiency and report quality(8–10).

With the recent emergence of artificial intelligence (AI), automated echo reporting has been proposed to use deep learning (DL) models to generate diagnostic predictions and measurements to fill a pre-set report template(8,10,11). These frameworks focused on specific image processing tasks(8,11) rather than the report text, and are technically equivalent to generating individual findings. However, these frameworks were not designed to handle the high-level cognitive activity of synthesizing clinically relevant impressions from detailed findings(12). In practice, physicians usually spend a significant amount of time summarizing detailed findings to clinically relevant final impressions(13,14). While this task is crucial, it can be time-consuming and prone to errors(15).

The advance of large language models (LLM) marked an important milestone for the application of AI in healthcare to automate clinical information summarization(13,14,16) and expert-level question-answering(17). A major advantage of LLMs is the flexibility of input and output(18), as well as the capability of handling conversations and interaction with human experts(19). While similar functionality can be achieved through commercially available LLMs (e.g., ChatGPT; OpenAI, San Francisco, CA)(20), only a few healthcare institutions have integrated ChatGPT(21). Furthermore, fine-tuning ChatGPT for specific tasks still requires uploading data to a central server, which also raises privacy concerns(22). In contrast, open-source LLMs are free of charge and can be locally fine-tuned for specific tasks within each healthcare institution’s secure confines(18).

Previous studies predominantly focused on electronic health records(13,16) and chest X-rays (CXR)(13,18) have highlighted the potential of using LLMs to summarize clinical text. In contrast, echo-related studies were mainly on data extraction or classification, rather than report summarization(23–25). Tang et al. used rule-based systems and a fine-tuned BART (Bidirectional and Auto-Regressive Transformer) model, EchoGen (26) for this purpose and demonstrated convincing results. However, EchoGen reports were less favored by human experts more than 50% of the time, perhaps due to the smaller number of parameters than current state-of-the-art LLMs(26). Meanwhile, despite a recent study exploring the use of an LLM for its question-answering capabilities on echo report texts(27), the potential of using billion-parameter LLMs to generate echo reports remains under-explored(28,29).

In this work, we proposed to evaluate LLMs in echocardiography reporting and construct a local, domain-specific LLM (EchoGPT) dedicated to echocardiography report summarization through an instruction fine-tuning approach, which is known to be an effective strategy to adapt LLMs for similar tasks(13,18). We anticipate that the fine-tuning procedure improves LLMs’ performance on the task of echocardiography report summarization. In addition to provide new insights into the use of open-source LLMs within the domain of echocardiography reporting, we will explore the challenges associated with applying current evaluation standards to echo reports generated by these models.

## Method

### Dataset

Mayo Clinic Reports: All adult (> 18 years old) echocardiography studies performed from 1/1/2017 to 12/31/2017 at Mayo Clinic Enterprise were retrieved. The types of studies include transthoracic echocardiography (TTE), transesophageal echocardiography (TEE), and stress echocardiography (including exercise and pharmacological studies). Text in the “Findings” and the “Final Impression” sections of each report was used for the current study (**Figure 1**). The study was approved by the Mayo Clinic IRB (protocol#: 22-010944).

**Figure 1.**
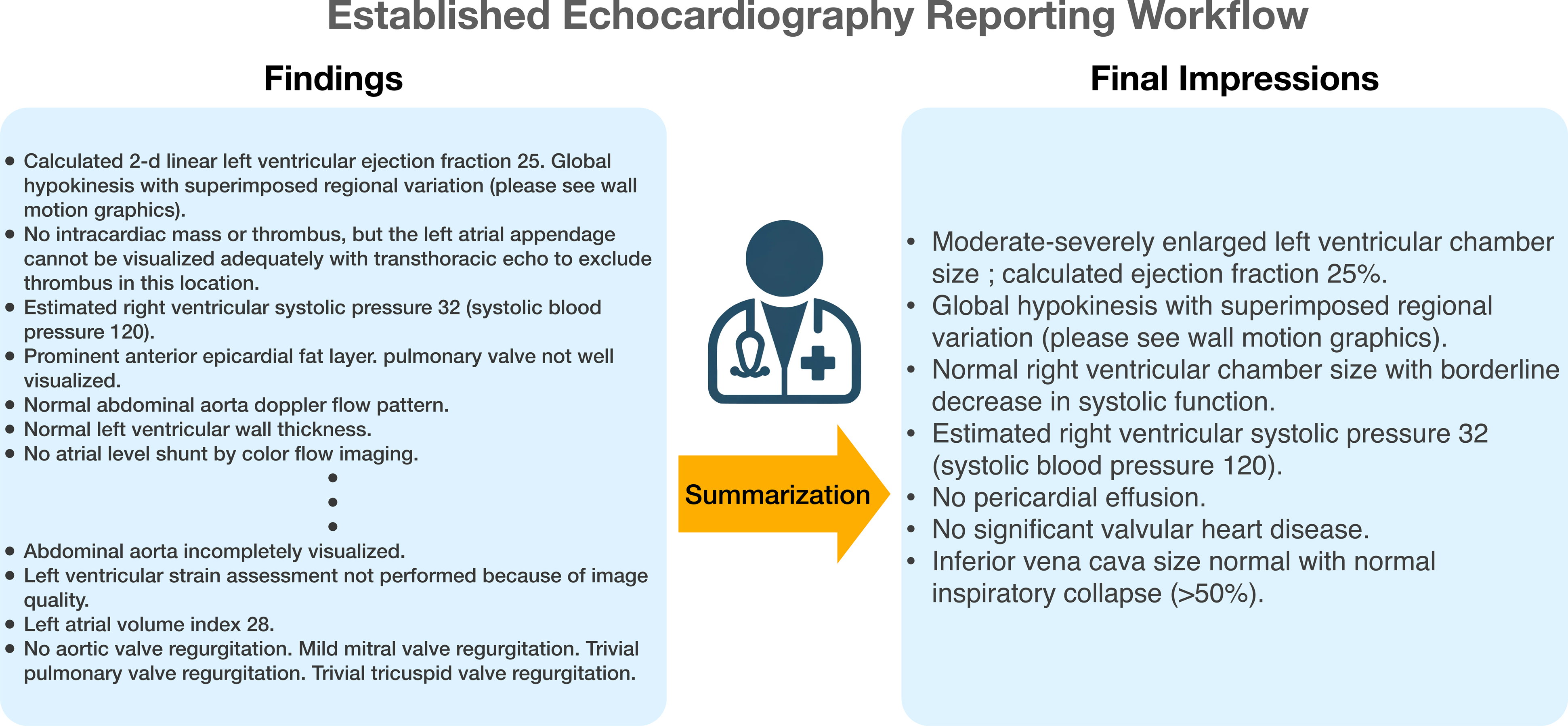
The current workflow of summarizing echo Findings into clinically relevant Final Impressions.

**Figure 2.**
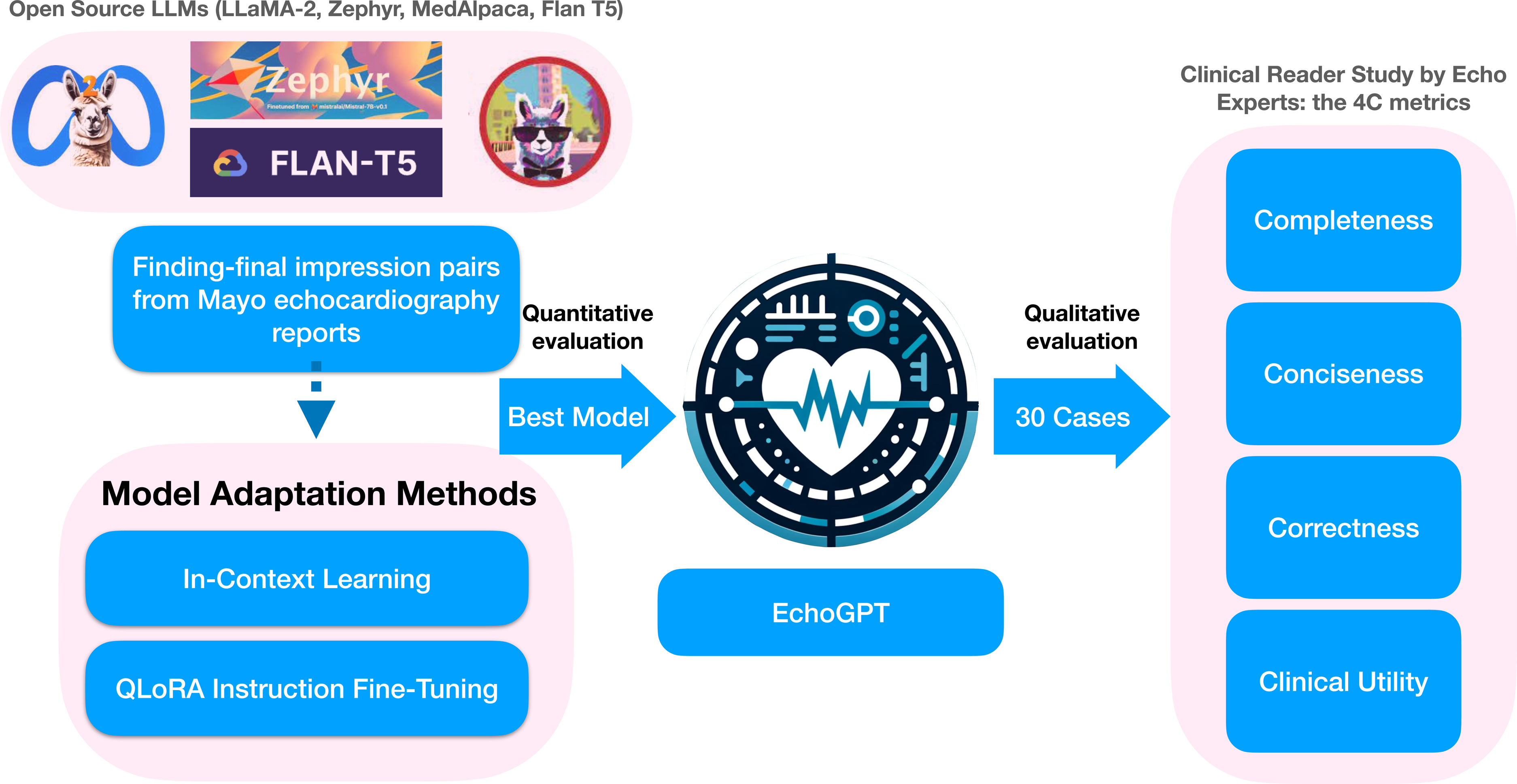
Overview of the EchoGPT study.

MIMIC-III ECHO-NOTE2NUM Dataset (v.1.0.0, referred to as MIMIC-EchoNotes below)(30): This publicly available dataset contains 43,472 valid free-text echocardiography reports from the intensive care unit at the Beth Israel Deaconess Medical Center between 2001 and 2012. A random subset of the dataset was used for external validation.

### Data Curation and Preprocessing

The summarization task was defined as creating the Final Impression section based on the Findings section, mirroring the established workflow of clinical echocardiography reporting at Mayo Clinic. The Final Impression text was used as the ground truth report (**Figure 1**).

Mayo Clinic Reports: Echocardiography reports were excluded according to the following criteria: (1) reports without Findings or Final Impression sections, (2) reports whose Findings or Impression section contained less than 15 words, as these are frequently canceled studies in our practice, and (3) labeled in report metadata as limited report. After this filtering process, the Findings section of each report was further processed as follows: (1) Remove capitalized subheadings (e.g., LEFT VENTRICLE, VALVES, OTHER FINDINGS, etc.), (2) Remove template sentences that make comparisons to prior reports, as no information from previous reports has been provided in Findings, and (3) Remove quality control-related sentences such as “study performed per left ventricular function protocol.”

MIMIC-EchoNotes Reports: Cases in this dataset were excluded based on criteria (1) and (2) above, as the metadata differed from that of the Mayo Reports. We also removed the subheadings and template sentences as described previously. We observed fundamental differences in report structure, including the “General Comments” section, which typically contains comments related to study quality, and the “Conclusions” section, which usually consists of the physician’s interpretation of findings. However, the Impression section often contains only 2-3 sentences summarizing the most pertinent study findings, which was challenging for head-to-head comparison in this study. Given the distinct report structure, the contents under the subheadings “General Comments” and “Conclusions” were integrated into the “Findings” and “Impression” sections, respectively. Common abbreviations in the text were expanded to their full forms.

### Data split

Data from Rochester, Florida, and Mayo Clinic Healthcare sites were used as the model development set. Considering variations in practice style among different sites, the data from the Arizona site was designated as the external validation set (referred to as the AZ validation set). Within the development set, 1,000 non-duplicated cases were randomly selected for the test and validation sets, respectively; the rest of the cases were used for fine-tuning (training set). Similarly, from the AZ validation and the MIMIC-EchoNotes datasets, we selected 1,000 non-duplicated random cases from each. For basic dataset statistics, the token length was calculated based on the natural language processing toolkit (NLTK) tokenizer(31), and the lexical variance was defined as the ratio of the number of unique tokens to the number of total tokens in each example(13).

### Model Selection

Due to patient privacy policy regulations, proprietary LLMs such as GPT-3.5 and GPT-4 were not considered in this work because versions of those models that were safe for protected health information were not yet available. For our target task, we selected auto-regressive and sequence-to-sequence LLMs with architectures under 7 billion parameters, balancing performance with manageability. Among open-source models, we selected representative auto-regressive models including Llama-2-7b-chat(28), Zephyr-7b(29), and Med-Alpaca(32) models considering their performance and max input context length on general natural language processing (NLP) tasks and radiology report summarization(13). For sequence-to-sequence (seq2seq) models, we used Flan-T5 (base) as the representative model as it is known for accurate text summarization(13,33), given that the EchoGen model(26) was not publicly available.

### Model Inference Hyperparameter Search

LLM inference was conducted by using Hugging Face’s (Manhattan, NY) transformer pipeline via the open-source LangChain framework(34). After initial tests, text generation and summarization were used as the task type for auto-regressive models and seq2seq models, respectively. A subset (10%, n=100) of examples were randomly selected from the test set for the hyperparameter search. We specifically tested the following configuration parameters that can significantly affect performance: temperature (0.1, 0.5, and 0.9) and repetition penalty (1.1, 1.2, and 1.3). These two parameters were tested separately, when one parameter was being tested, the other was fixed at the lowest value. The generated contents were evaluated by both automatic metrics and qualitative assessment. We chose the following configuration for model inference: {temperature 0.1, repetition penalty 1.1} after comparing automatic metrics and qualitative assessments; see **Supplemental Table 1**). We did not complete a dedicated search procedure for the optimal LLM inference configurations, but the configurations used in our study were similar to prior reports, and the generated contents were satisfying on qualitative review. Of note, the configurations were tested in a zero-shot setting, and the best configuration was directly applied to the ICL and QLoRA fine-tuned models(13).

### Model Adaptation

#### Prompt

A prompt template was created with components of the prefix, instruction, and suffix(13) (**Table 1**). The final prompt was decided after qualitatively evaluating several different variants of each component on a small subset of the data. We also specified that the summarization should be “concise” and use “a minimal amount of text” to avoid LLMs generating lengthy reports(13). Likely due to the difference in the reporting style of the MIMIC-EchoNotes dataset, the final prompt above led to suboptimal responses. Therefore, we adopted a new prompt tailored to match the reporting style by incorporating new instructions below: 1) Write a 10-bullet points clinical summary, and 2) Avoid using numbers other than LVEF.

**Table 1.** Prompt Template.

|  | Prompt Component |
| --- | --- |
| Prefix | “You are a knowledgeable cardiologist.” |
| Instruction | “For the following echocardiography report findings, please write a concise summary with a minimal amount of text.” |
| Suffix (ICL only) | “Use the following examples to guide word choice.” |

#### In-context Learning (ICL)

ICL has been proposed to improve LLM’s performance without changing the base model weights(13,35,36). Also, using relevant in-context examples is shown to have better model performance compared to random examples in ICL. To obtain relevant in-context examples, we adopted the approach to select m (m=1, 2, 4…) nearest neighbors from the training set for each test set case, after embedding both sets by the PubMedBERT model(37).

#### Instruction tuning with quantized low-rank adaptation (QLoRA)

Due to the size of candidate models, we opted for quantized low-rank adaptation (QLoRA)(38), a type of parameter efficient fine-tuning (PEFT)(39) to optimize our LLMs for echo report summarization tasks. The same prompt template (**Table 1**) was used, and the Final Impression text from the same report was used as the target output (13,14).

We configured the training process as follows: load model in 4-bit precision, with a LoRA configuration of (alpha = 16, LoRA dropout = 0.1, LoRA r= 64). The batch size and gradient accumulation were adjusted for each model to achieve an effective batch size of 24 that fits on a single NVIDIA RTX A5000 24G GPU setting. A paged-AdamW 32-bit optimizer was used, with an initial learning rate of 1e-3, which decayed to 1e-4 (by a cosine scheduler) after the initial 100 warm-up steps. The above configuration provided the most stable training process after attempting different configurations reported in prior studies(13,38).

### Model Performance Evaluation

#### Automatic NLP evaluation metrics

To evaluate the models’ performance on the information summarization task and compare it to prior works, we utilized four established automatic metrics that have been used in other clinical text summarization studies(40,41): BLEU (Bilingual Evaluation Understudy)(42), METEOR (Metric for Evaluation of Translation with Explicit ORdering)(43), ROUGE-L (Recall-Oriented Understudy for Gisting Evaluation - Longest Common Subsequence), and the BERT (Bidirectional Encoder Representations from Transformers) score(44), which represents the similarity between generated contents and the corresponding reference at words/characters (n-gram), single word (unigram), longest sequence of words, and contextual level, respectively For ROUGE-L, we present the F1 score component(26,45). For factual correctness, the RadGraph-F1 metric (level: all) was reported(46,47) This metric served as the primary evaluation criterion for model performance, considering its significance in ensuring the factual correctness of generated clinical content.

#### Evaluation of Significance of Measurement Numbers in Automatic Metrics

Considering the importance of measurements in echo studies, we also attempted to evaluate whether the current automatic metrics can detect changes in measurement numbers. For this purpose, we generated synthetic reports by replacing all the measurement numbers with random numbers ranging from 1 to 99 in the reports. We then compared the automatic metric scores with the corresponding reports with the original measurements.

#### Human expert evaluation metrics

We designed a human expert evaluation process based on previous clinical text summarization studies(13,18,26). The Findings with corresponding ground truth (Final Impression) and LLM-generated summarization of 30 randomly selected cases were presented to four echocardiography-board-certified cardiologists for blinded quality review. We noted that physician-summarized Impressions may contain free-text information beyond the Findings (e.g., documenting events during the study or communication with the ordering provider), and the reviewers were instructed to rate only based on information within the Findings section. Each metric was rated for the preference (5 levels) between the two summarizations (**Supplemental Figure 1**)(13). The “4C metrics” evaluated by echocardiography experts are completeness, conciseness, correctness, and clinical utility(14), as described in **Table 2**.

**Table 2.**
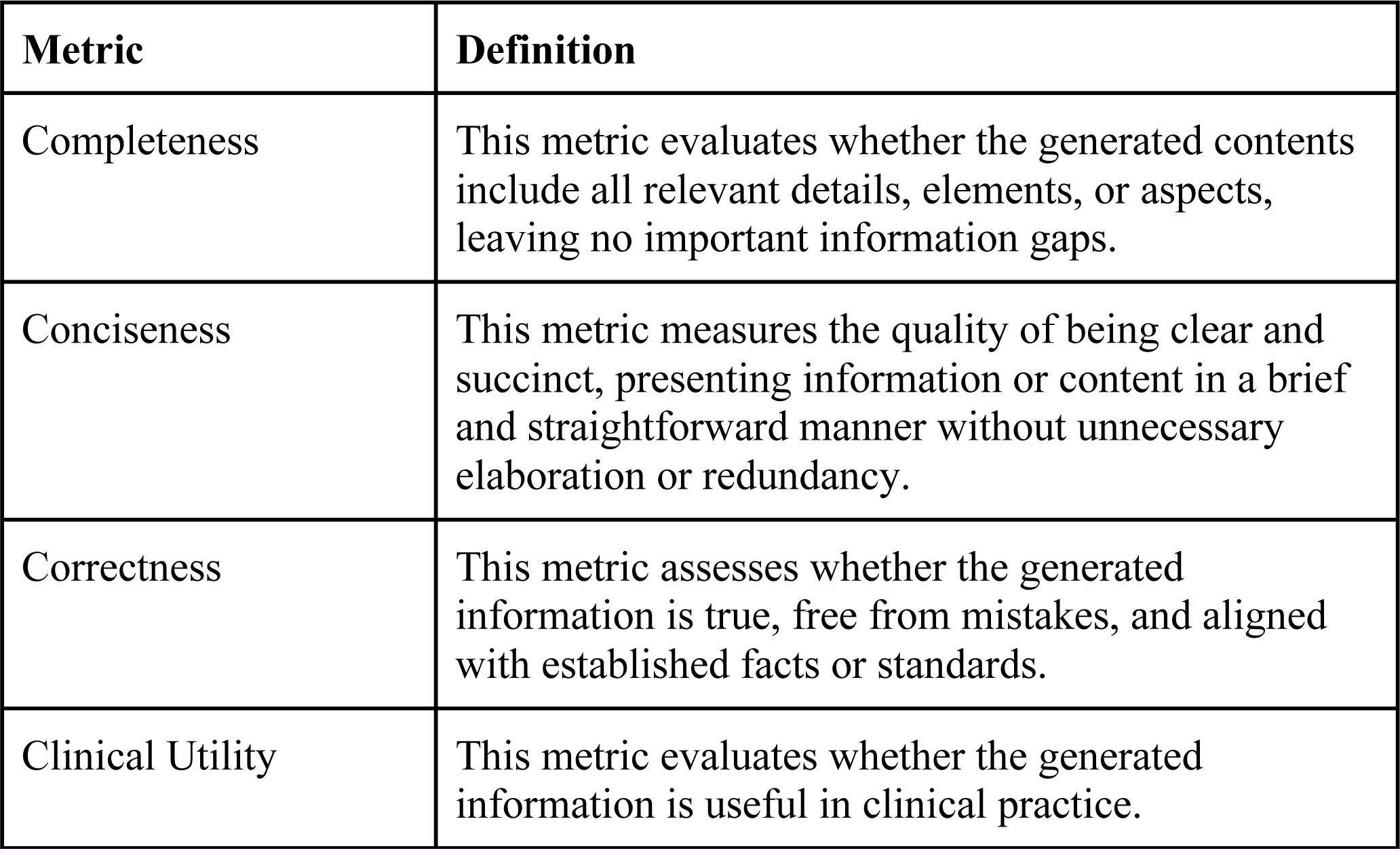
Definition of the 4C Human Expert Evaluation Metrics.

| Metric | Definition |
| --- | --- |
| Completeness | This metric evaluates whether the generated contents include all relevant details, elements, or aspects, leaving no important information gaps. |
| Conciseness | This metric measures the quality of being clear and succinct, presenting information or content in a brief and straightforward manner without unnecessary elaboration or redundancy. |
| Correctness | This metric assesses whether the generated information is true, free from mistakes, and aligned with established facts or standards. |
| Clinical Utility | This metric evaluates whether the generated information is useful in clinical practice. |

#### Statistical analysis

Automatic metric performance between each pair of models was compared using a two-tailed paired Student’s t-test or Wilcoxon signed-rank test for normally and non-normally distributed data, respectively. Models were also compared based on win rates, which are defined as the percentage of head-to-head victories in performance between two models for each selected metric(13). For the performance bias analysis, data were grouped based on sex (male versus female) and race (white, black, other). In the case of sex, we employed two-tailed Student’s t-tests or Mann-Whitney U tests for normally and non-normally distributed data, respectively. For race, we utilized one-way ANOVA. In human expert qualitative analysis, the 4C metrics were compared by a one-sample Wilcoxon signed-rank test(13), and the agreement of ratings between experts was assessed by Fleiss’ kappa coefficient(48), and interpreted as recommended by Landis et al(49). We conducted Pearson correlation analyses to explore the relationships between each human expert evaluation metric, assessing their independence. Additionally, we conducted similar analyses to examine the correlation between human and automatic metrics. Statistical analyses were performed using Python 3.8 and SciPy 1.8.0. All the comparisons consider a p-value < 0.05 as significant.

## Results

### Patient Cohort and Dataset

Our development set contains 97,506 reports from 71,717 unique patients, with a mean age of 64.3±15.8 years, 54,005 (55.4%) were male, 89,466 (91.8%) were white. Randomly selected from Mayo Arizona studies (19,557 reports/15,853 unique patients), the AZ validation set contains 1,000 reports from 1,000 unique patients with a mean age of 63.9 ±16.0 years, 584 (58.4%) were male and 885 (88.5%) were white. Detailed demographic information was not available for the MIMIC-EchoNotes dataset. Other detailed patient characteristics and statistics of text data are summarized in **Table 3**. Transthoracic echocardiography was the predominant study type in development, AZ validation, and MIMIC-Echo datasets (81.9%,77.6%, and 86.4% respectively).

**Table 3.** Data distribution of the development and AZ validation sets.

|  | Development Set |  |  |  | AZ validation set | MIMIC-EchoNotes |
| --- | --- | --- | --- | --- | --- | --- |
|  | All | Train | Validation | Test |  |  |
|  | n= 97,506 | n= 95,506 | n=1,000 | n=1,000 | n=1,000 | n=1,000 |
| Age | 64.3 ± 15.8 | 64.3 ± 15.8 | 64.9 ± 16.0 | 64.7 ± 15.7 | 63.9 ± 16.0 | -- |
| Race |  |  |  |  |  |  |
| *White* | 89466 (91.8%) | 87632 (91.8%) | 914 (91.4%) | 920 (92.0%) | 885 (88.5%) | -- |
| *Black* | 3326 (3.4%) | 3257 (3.4%) | 35 (3.5%) | 34 (3.4%) | 42 (4.2%) | -- |
| *Other* | 2850 (2.9%) | 2790 (2.9%) | 28 (2.8%) | 32 (3.2%) | 35 (3.5%) | -- |
| *Asian* | 1415 (1.5%) | 1390 (1.5%) | 16 (1.6%) | 9 (0.9%) | 22 (2.2%) | -- |
| *Native American* | 362 (0.4%) | 353 (0.4%) | 5 (0.5%) | 4 (0.4%) | 13 (1.3%) | -- |
| *Pacific Islander* | 87 (0.1%) | 84 (0.1%) | 2 (0.2%) | 1 (0.1%) | 3 (0.3%) | -- |
| Sex |  |  |  |  |  |  |
| *Male* | 54010 (55.4%) | 52920 (55.4%) | 563 (56.3%) | 527 (52.7%) | 584 (58.4%) | -- |
| *Female* | 43496 (44.6%) | 42586 (44.6%) | 437 (43.7%) | 473 (47.3%) | 416 (41.6%) | -- |
| HTN | 11578 (11.9%) | 11320 (11.9%) | 115 (11.5%) | 143 (14.3%) | 125 (12.5%) | -- |
| DM | 14406 (14.8%) | 14105 (14.8%) | 142 (14.2%) | 159 (15.9%) | 115 (11.5%) | -- |
| CAD | 4983 (5.1%) | 4863 (5.1%) | 51 (5.1%) | 69 (6.9%) | 57 (5.7%) | -- |
| CHF | 38908 (39.9%) | 38105 (39.9%) | 384 (38.4%) | 419 (41.9%) | 293 (29.3%) | -- |
| CKD | 15071 (15.5%) | 14759 (15.5%) | 140 (14.0%) | 172 (17.2%) | 170 (17.0%) | -- |
| Stroke | 2879 (3.0%) | 2812 (2.9%) | 30 (3.0%) | 37 (3.7%) | 21 (2.1%) | -- |
| Echo Study Type |  |  |  |  |  |  |
| *Adult TTE* | 79902 (81.9%) | 78243 (81.9%) | 833 (83.3%) | 826 (82.6%) | 776 (77.6%) | 881 (88.1%) |
| *Adult TEE* | 7829 (8.0%) | 7685 (8.0%) | 65 (6.5%) | 79 (7.9%) | 63 (6.3%) | 108 (10.8%) |
| *Exercise Stress* | 6781 (7.0%) | 6652 (7.0%) | 64 (6.4%) | 65 (6.5%) | 131 (13.1%) | 11 (1.1%) |
| *Pharmacological Stress* | 2994 (3.1%) | 2926 (3.1%) | 38 (3.8%) | 30 (3.0%) | 30 (3.0%) | -- |
| Data Characteristics |  |  |  |  |  |  |
| *Average Number of Tokens (Findings)* | 215.7 ± 55.8 | 215.7 ± 55.9 | 213.1 ± 55.2 | 214.8 ± 53.3 | 217.1 ± 77.3 | 184.3 ± 50.0 |
| *Average Number of Tokens (Final Impression)* | 88.2 ± 34.3 | 88.2 ± 34.3 | 87.1 ± 33.7 | 86.8 ± 33.6 | 97.7 ± 37.9 | 163.2 ± 40.4 |
| *Average Lexical Variance (Findings)* | 0.51 | 0.51 | 0.52 | 0.51 | 0.52 | 0.54 |
| *Average Lexical Variance (Final Impression)* | 0.65 | 0.65 | 0.65 | 0.65 | 0.64 | 0.52 |
| CAD: coronary artery disease, HTN: hypertension, DM: diabetes, CKD: chronic kidney disease, CHF: congestive heart failure. TTE: transthoracic echocardiography, TEE: transesophageal echocardiography. |  |  |  |  |  |  |

### Zero-shot, ICL, and QLoRA fine-tuned performance

**Table 4** is a summary of the performance of zero-shot and fine-tuned LLMs, including MedAlpaca, Llama-2, and Zephyr. QLoRA fine-tuning significantly improved LLMs’ performance from baseline. Note that T5 and MedAlpaca were not fine-tuned so only zero-shot results were provided for reference. Among the candidate models, Llama-2 generally had the best zero-shot performance, which was consistent in ICL. While Flan-T5 had a similar or superior performance to Llama-2 across most metrics, it was particularly worse on the RadGraph F1 score (**Table 4**; **Figure 3**). On qualitative review, we noted that T5 provided concise summaries, however important clinical information was missed in this process (**Supplemental Table 2**).

**Figure 3.**
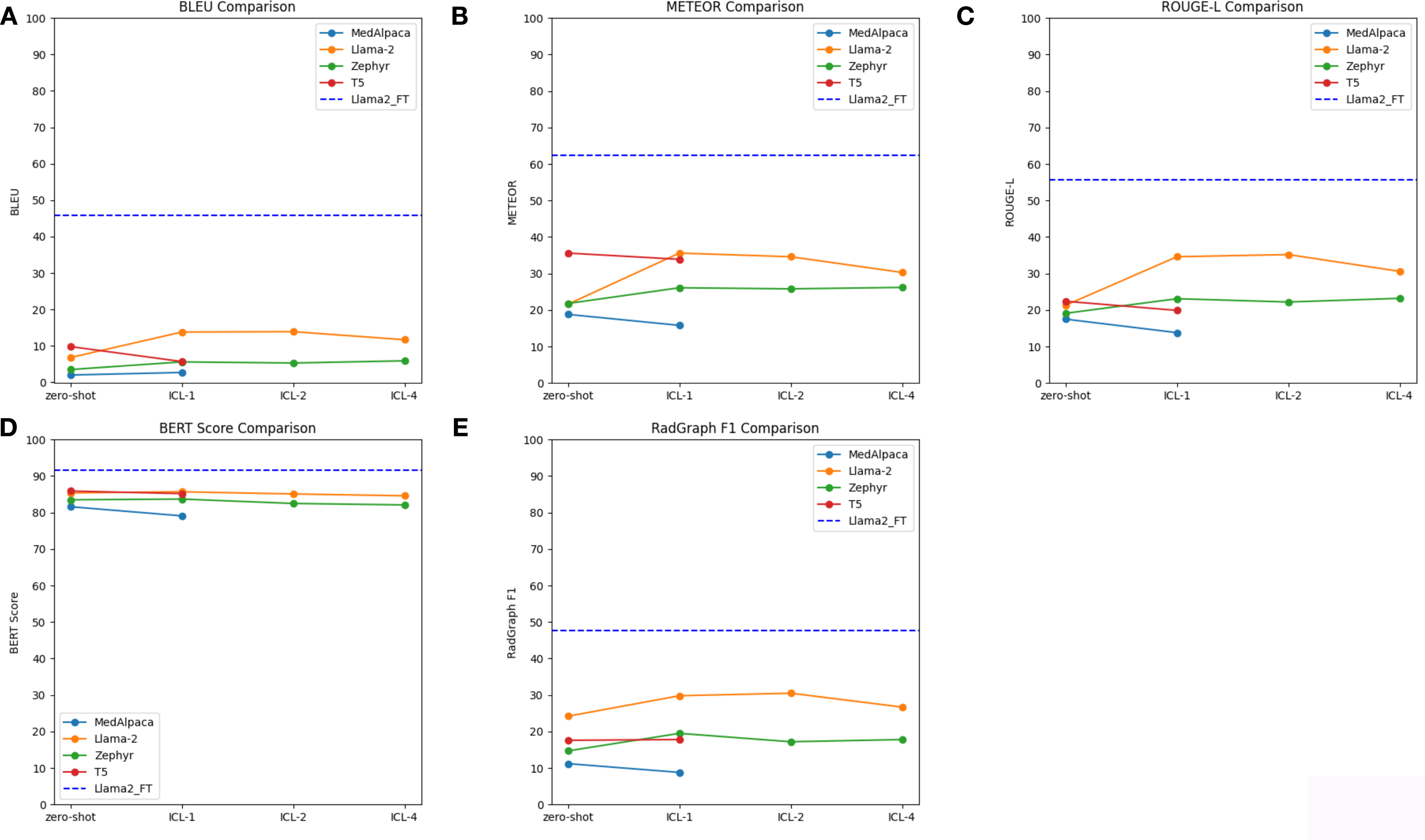
ICL performance of each LLM. Panel A to E correspond to BLEU, METEOR, ROUGE-L, BERT Score, and RadGraph F1 Score, respectively. Zero-shot and fine-tuned Llama-2 (EchoGPT; horizontal purple dashed line) performance was included for reference.

**Table 4.**
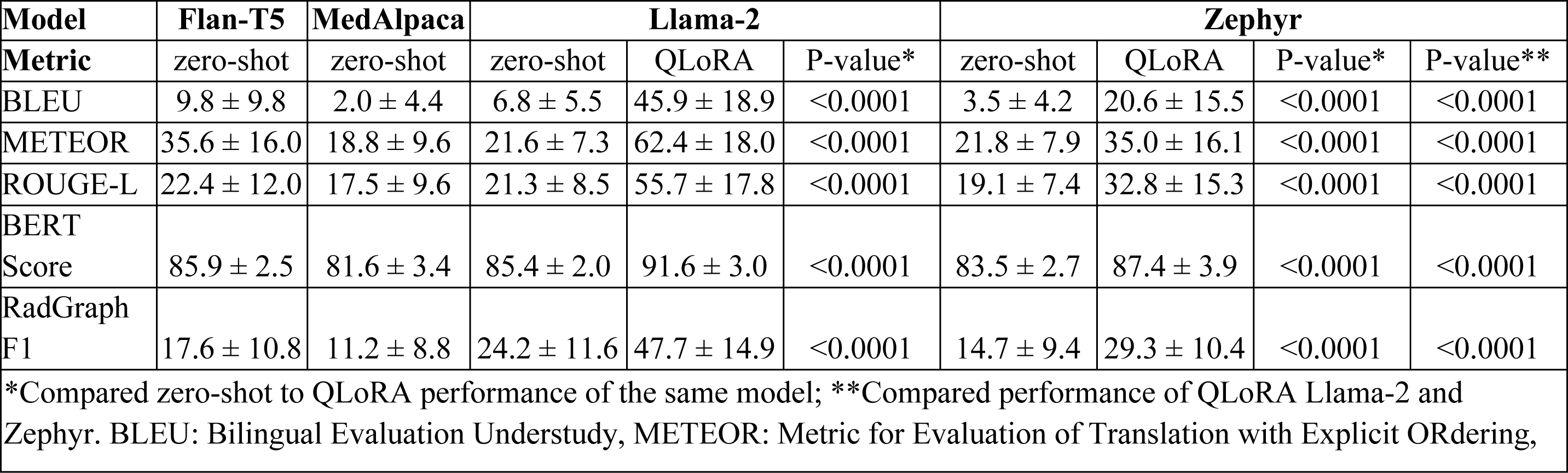
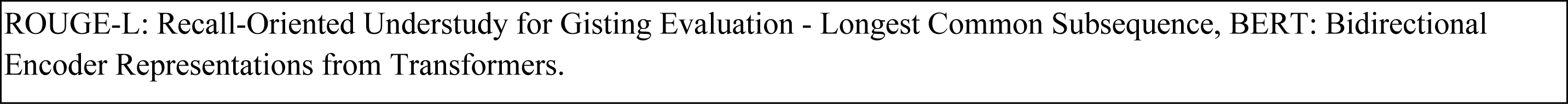
Quantitative performance of zero-shot and QLoRA fine-tuned LLMs.

| Model | Flan-T5 | MedAlpaca | Llama-2 |  |  | Zephyr |  |  |  |
| --- | --- | --- | --- | --- | --- | --- | --- | --- | --- |
| Metric | zero-shot | zero-shot | zero-shot | QLoRA | P-value* | zero-shot | QLoRA | P-value* | P-value** |
| BLEU | 9.8 ± 9.8 | 2.0 ± 4.4 | 6.8 ± 5.5 | 45.9 ± 18.9 | <0.0001 | 3.5 ± 4.2 | 20.6 ± 15.5 | <0.0001 | <0.0001 |
| METEOR | 35.6 ± 16.0 | 18.8 ± 9.6 | 21.6 ± 7.3 | 62.4 ± 18.0 | <0.0001 | 21.8 ± 7.9 | 35.0 ± 16.1 | <0.0001 | <0.0001 |
| ROUGE-L | 22.4 ± 12.0 | 17.5 ± 9.6 | 21.3 ± 8.5 | 55.7 ± 17.8 | <0.0001 | 19.1 ± 7.4 | 32.8 ± 15.3 | <0.0001 | <0.0001 |
| BERT Score | 85.9 ± 2.5 | 81.6 ± 3.4 | 85.4 ± 2.0 | 91.6 ± 3.0 | <0.0001 | 83.5 ± 2.7 | 87.4 ± 3.9 | <0.0001 | <0.0001 |
| RadGraph F1 | 17.6 ± 10.8 | 11.2 ± 8.8 | 24.2 ± 11.6 | 47.7 ± 14.9 | <0.0001 | 14.7 ± 9.4 | 29.3 ± 10.4 | <0.0001 | <0.0001 |
| *Compared zero-shot to QLoRA performance of the same model; **Compared performance of QLoRA Llama-2 and Zephyr. BLEU: Bilingual Evaluation Understudy, METEOR: Metric for Evaluation of Translation with Explicit ORDERing, |  |  |  |  |  |  |  |  |  |

ROUGE-L: Recall-Oriented Understudy for Gisting Evaluation - Longest Common Subsequence, BERT: Bidirectional Encoder Representations from Transformers.

In ICL, LLMs that allow longer context length (Llama-2 and Zephyr) had the best performance across all metrics when one example was provided (ICL-1). The performance gradually trended down with more examples (ICL-2 and ICL-4). In contrast, the performance of LLMs with shorter max context length started to trend down with one example (**Figure 3**). Importantly, we observed that Llama-2 can integrate information from multiple ICL examples, such as “Calculated 2-d linear left ventricular ejection fraction 57, 61, 75 (3 reports)”, which occurred in 34 (3.4%) and 69 (6.9%) cases for ICL-2 and ICL-4, respectively (**Supplemental Table 3**).

Based on the zero-shot and ICL performance of the candidate models, Llama-2 and Zephyr were selected for instruction fine-tuning. Compared to zero-shot, QLoRA significantly improved the performance of selected LLMs across all metrics (**Table 4**). For the head-to-head comparison in model win rates, fine-tuned Llama-2 was superior to all other models, including Zephyr (base and fine-tuned), MedAlpaca (base), and Flan-T5 (base) across all 5 automatic metrics (**Figure 4**). Llama-2 maintained similar performance in the AZ validation set (n=1,000) and was consistently superior to fine-tuned Zephyr (**Supplemental Table 4**). Regarding potential biases, we did not observe significant biases regarding sex and race across the automatic metrics, except for a slightly better RadGraph F1 performance in female patients in the AZ validation set (male vs. female: 0.38 ± 0.14 vs. 0.40 ± 0.15, p=0.04) (**Supplemental Figure 2**). Because Llama-2 had the best performance on zero-shot, ICL, and QLoRA fine-tuning approaches, fine-tuned Llama-2 was selected as EchoGPT and used for the subsequent expert qualitative review.

**Figure 4.**
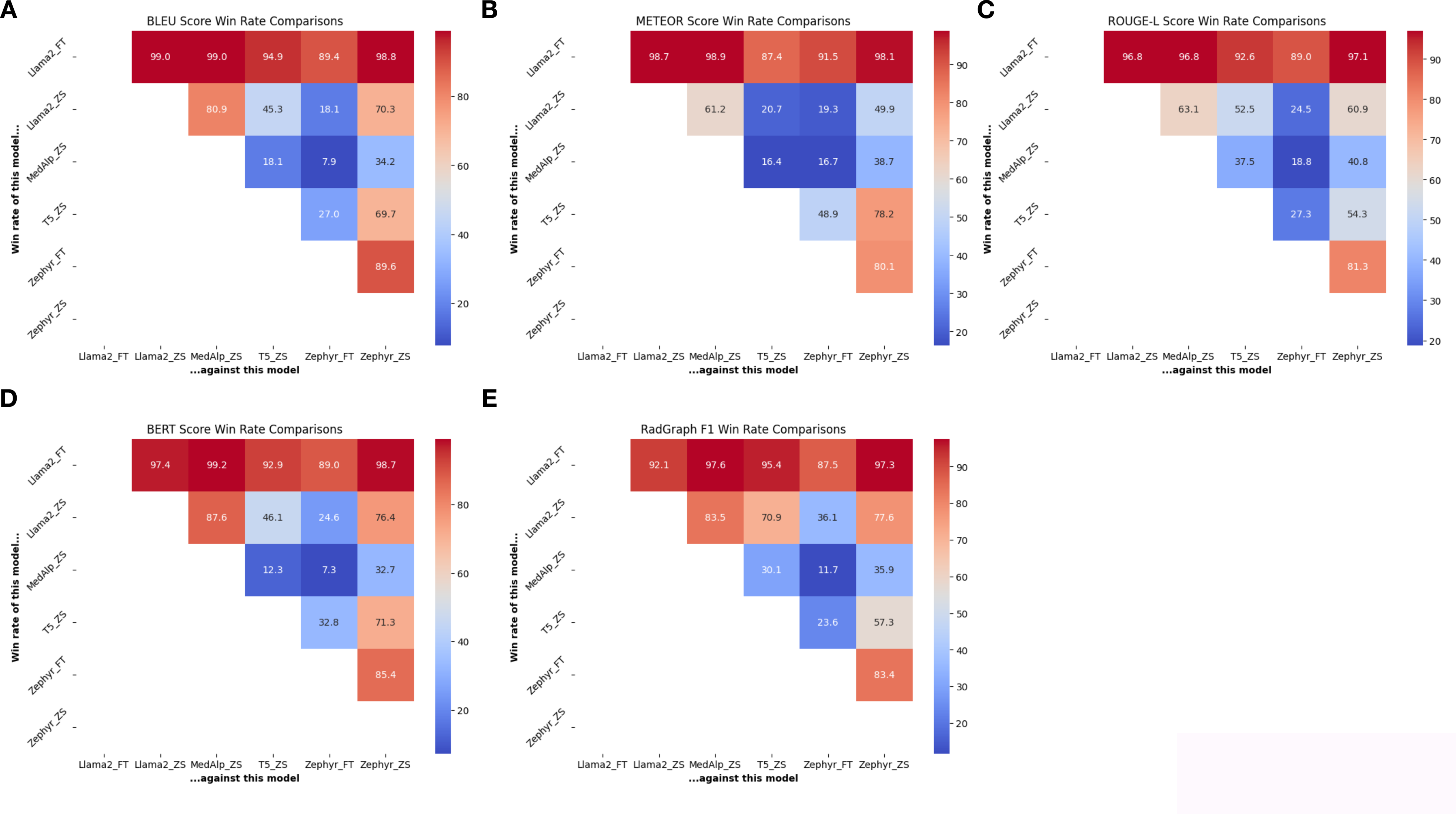
Model win rates on the test set. Model win rate heatmap illustrates the head-to-head win rate comparisons (in percentile) among different models based on the selected metrics. Cool colors indicate lower win rates and warmed colors indicate higher win rates. We compared Llama-2 (base and fine-tuned), Zephyr (base and fine-tuned), T5 (base), and MedAlpaca (base). Fine-tuned Llama-2 consistently outperformed all other models across all 5 automatic metrics. FT: fine-tuned. ZS: zero-shot (base model).

### Significance of Measurement Numbers in Automatic Metrics

After replacing the measurement numbers with random numbers, we observed relatively minor decreases in BLEU, METEOR, ROUGE-L, BERT, and RadGraph F1 scores, while statistically significant (p<0.0001). One can see that in the provided examples, the report with random numbers doesn’t make clinical sense when compared to the original content (**Table 5**).

**Table 5.**
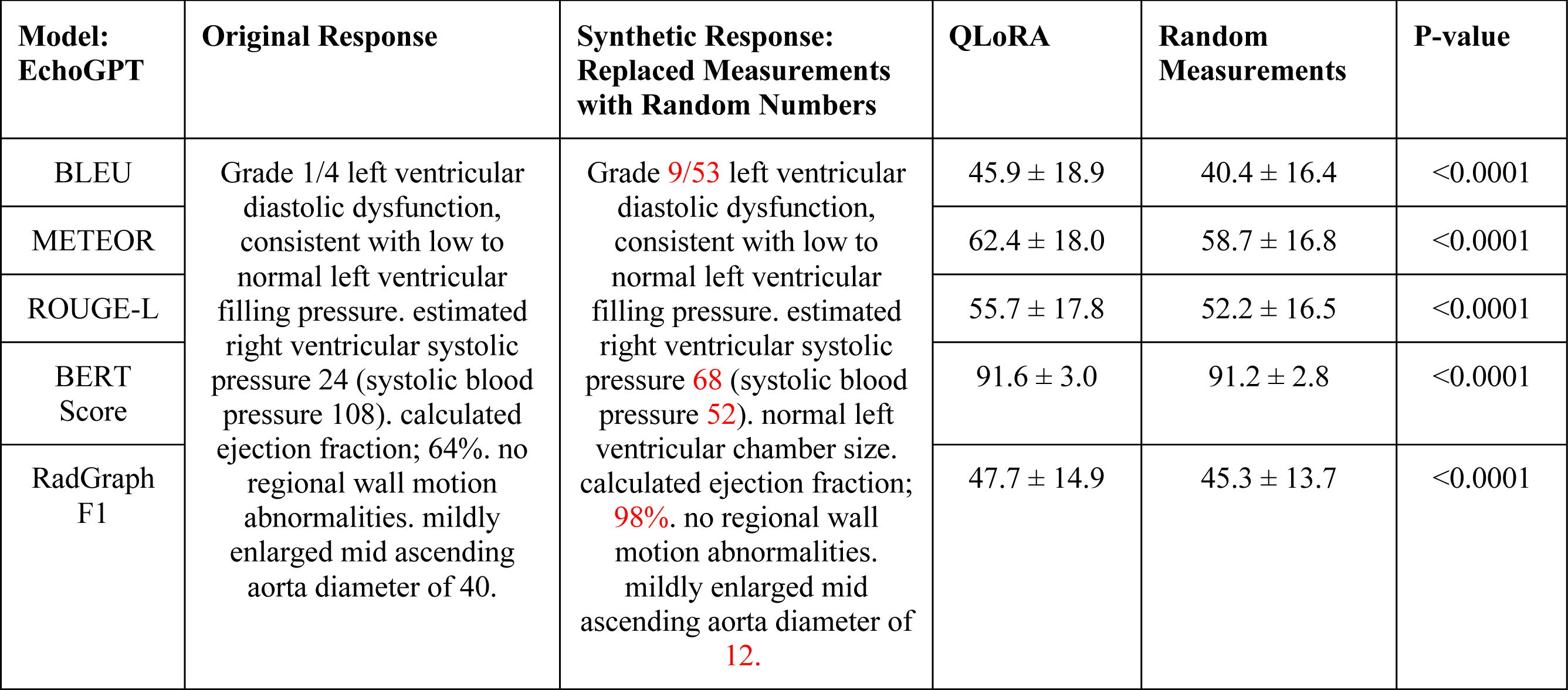
Comparison of Automatic Metrics between the Original Response and the Synthetic Response.

### Human Expert Evaluation

We observed slight agreement for correctness, fair agreement for conciseness and clinical utility, and moderate agreement for completeness (**Supplemental Table 5**). Among the 4C metrics, we observed that EchoGPT significantly outperformed human experts in conciseness (p<0.001). There was no significant preference among the other three categories (**Figure 5A**). The 4C metrics were not completely independent. There was a high correlation between clinical utility and completeness (Pearson’s r= 0.78), and modest to moderate correlations between other metrics (**Figure 5C**). We also observed that across all automatic metrics, RadGraph F1 had modest to moderate correlations with all 4 human evaluation metrics (**Figure 5C**).

**Figure 5.**
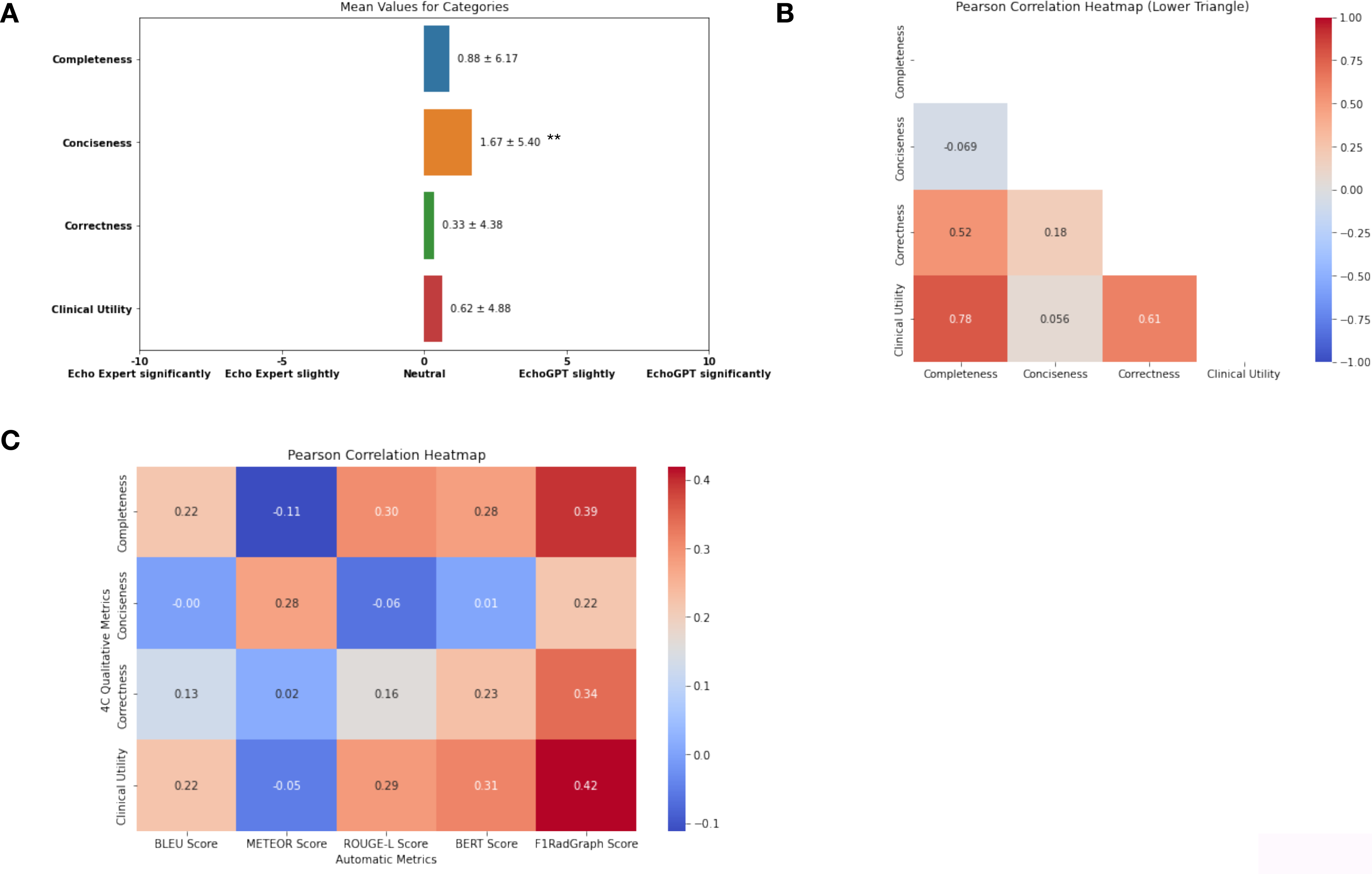
Human expert qualitative evaluation results. **Panel A**. In the 4 categories, EchoGPT significantly outperformed human experts in conciseness (p<0.001). We didn’t observe significant differences among the other three categories (completeness, correctness, and clinical utility). **Panel B.** showed interdependence of the 4C metrics, especially the correlations between clinical utility and completeness (Pearson’s r= 0.78), and modest to moderate correlations between other metrics. **Panel C.** Correlations between automatic metrics and the 4C metrics. Across all automatic metrics, RadGraph F1 had modest to moderate correlations with all 4 human evaluation metrics. Preference ratings were expressed as mean ± standard deviation, **indicates p<0.001.

### External Validation on the MIMIC-EchoNotes dataset

In the MIMIC-EchoNotes dataset (n=1,000), we observed a performance drop in fine-tuned models, while EchoGPT was still superior to fine-tuned Zephyr (**Supplemental Table 4**). Regarding the expert review, we observed moderate agreement for completeness and conciseness, but slight agreement for correctness and clinical utility (**Table 6**). The original reports (combined Conclusions and Impression sections) were preferred over EchoGPT in completeness, correctness, and clinical utility (p< 0.001); while EchoGPT was superior in conciseness (p<0.001) (**Supplemental Figure 6A**). RadGraph F1 still had the strongest correlations with the 4C metrics, although other metrics also had stronger correlations (**Supplemental Figure 3**). A representative example demonstrated the difference in reporting structure and style, along with reviewers’ feedback on the two datasets (**Table 6**).

**Table 6.** Representative Examples in Reader Study.

| Mayo Echo<br>Report | Findings | Impression | EchoGPT Summary |
| --- | --- | --- | --- |
| **Representative Example** | Calculated 2-d linear left ventricular ejection fraction 61. No regional wall motion abnormalities. aortic valve strands (on the ventricular side of the valve). Estimated right ventricular systolic pressure 23 (systolic blood pressure 108). Normal abdominal aorta doppler flow pattern. Normal left ventricular wall thickness. No atrial level shunt by color flow imaging. left atrial volume index 30. Trivial aortic valve regurgitation. Trivial mitral valve regurgitation. Trivial pulmonary valve regurgitation. Trivial tricuspid valve regurgitation. No pericardial effusion. Normal aortic valve. thickness. Normal pulmonary valve. Normal mitral valve. Normal tricuspid valve. Trileaflet aortic valve. Normal left ventricular chamber size. Normal right ventricular chamber size. Normal right atrial size. Normal left atrial size with normal inspiratory collapse (>50%) inferior vena cava size normal. Normal ascending aorta diameter. Normal right ventricular systolic function. Normal pulmonary valve systolic velocities. No abdominal aortic aneurysm. Normal left ventricular diastolic function. Calculated 2-d monoplane volumetric left ventricular ejection fraction 59. | Normal left ventricular chamber size. Calculated ejection fraction 61%. No regional wall motion abnormalities. Normal right ventricular chamber size and function. Estimated right ventricular systolic pressure 23 mmHg. Inferior vena cava size: normal, with normal inspiratory collapse (>50%). Normal left ventricular diastolic function. | No regional wall motion abnormalities. Normal cardiac valves. Estimated right ventricular systolic pressure 23 (systolic blood pressure 108). No pericardial effusion. Normal left ventricular chamber size and wall thickness. Calculated left ventricular ejection fraction 61%. Normal right ventricular chamber size and systolic function. Inferior vena cava size: normal, with normal inspiratory collapse (>50%) |
| **Comments** | Numbers were utilized in pertinent statements, such as right ventricular systolic pressure, systolic blood pressure, ejection fraction, and left atrial volume index. Typical reports detail the method of LVEF measurement alongside the value. | This section typically contains a 7–10-point summary that covers the overall condition of the heart. In this case, no information beyond the Findings was included. | EchoGPT captured the relevant findings, although not in the same order. LV ejection fraction measurement was included in the summary (61%). |
| **MIMIC-EchoNote Report** | **Findings** | **Conclusions/Impression** | **EchoGPT Summary** |
| **Representative Example** | Highly mobile echodensity (0.8x0.5 cm) in the right atrium, best seen on right ventricle inflow views, which appears to be attached to a catheter (catheter not fully visualized, but patient with history of port-A-cath). Doppler across tricuspid valve suggestive of tricuspid valve prolapse. Normal left atrium size. Normal right atrium size. right atrium mass. Normal inferior vena cava diameter ($\leq 2.1$ cm) with $>50\%$ decrease with sniff (estimated right atrium pressure (0-5 mmHg). Normal left ventricle wall thickness, cavity size and regional/global systolic function (left ventricular ejection fraction $>55\%$). Normal right ventricle chamber size and free wall motion. Normal diameter of aorta at the sinus, ascending and arch levels. No 2D or Doppler evidence of distal arch coarctation. Normal aortic valve leaflets (3). No aortic stenosis. No aortic regurgitation. Normal mitral valve leaflets with trivial mitral regurgitation. TVP. Mild tricuspid regurgitation. Normal pulmonic valve leaflet. No PS. Physiologic pulmonary regurgitation. No pericardial effusion. If clinically indicated, a transesophageal echocardiographic examination is recommended. Echocardiographic results were reviewed by telephone with the MD caring for the patient. | The left atrium is normal in size. A highly mobile mass, measuring 0.8x0.5 cm, is seen in the right atrium (possibly attached to a catheter?) which may represent thrombus or vegetation. The estimated right atrial pressure is 0-5 mmHg. Left ventricular wall thickness, cavity size and regional/global systolic function are normal (left ventricular ejection fraction $>55\%$). Right ventricular chamber size and free wall motion are normal. The diameters of aorta at the sinus, ascending and arch levels are normal. The aortic valve leaflets (3) appear structurally normal with good leaflet excursion and no aortic stenosis or aortic regurgitation. Tricuspid valve prolapse is present based on doppler findings with mild tricuspid regurgitation. The mitral valve appears structurally normal with trivial mitral regurgitation. There is no pericardial effusion. **Highly mobile mass is seen in the right atrium (possibly attached to a catheter?) which may represent thrombus or vegetation. Clinical correlation advised. Normal biventricular systolic function. Tricuspid valve prolapse with mild tricuspid regurgitation. If clinically indicated, a transesophageal echocardiographic examination is recommended. Drs. and were notified of these findings at 4:45 p.m. today.** | Mass noted in right atrium, measuring 0.8 x 0.5cm, attached to a catheter (catheter not fully visualized but has history of port-A-cath). Doppler across tricuspid valve suggestive of tricuspid valve prolapse. Normal left ventricular chamber size, normal regional/global systolic function, estimated ejection fraction 60%. Normal right ventricular chamber size and function. Normal diameter of aorta at the sinus, ascending and arch levels. No 2D or Doppler evidence of distal arch coarctation. No pericardial effusion. If clinically indicated, a transesophageal echocardiographic examination is recommended. |
| **Comments** | In the findings section, normal LVEF was described as 'LVEF $> 55\%$' without further specification. Measurements such as right ventricular systolic pressure, and LA volume index were not included in the report. | The combined section was longer than the Findings section. The blue and red text belongs to the Conclusions and Impression sections, respectively. Information beyond the Findings was in the text: "Drs. and were notified..." | Relatively brief summary generated by EchoGPT, capturing the important findings (right atrial mass attached to catheter) and relevant recommendations. However, EchoGPT hallucinated estimated ejection fraction- which was a template sentence used at Mayo. |

## Discussion

This study evaluated the feasibility of multiple open-source LLMs in echocardiography report summarization through different model adaptation methods. Trained on one of the largest echocardiography report datasets in the world, we demonstrated that QLoRA fine-tuning can significantly improve LLMs’ performance for the desired summarization task, with at least comparable qualities to human experts. ICL was overall inferior to QLoRA fine-tuning and faced substantial limitations, such as integrating information from multiple examples. Additionally, we demonstrated that current automatic metrics are not sensitive to the change in measurement numbers in echo reports. The current study offers insights into the development and evaluation of a specialized local LLM tailored for echo report summarization and presenting significant workflow advantages.

### Model Adaption Approaches: ICL vs. Fine-tuning

Our results suggested that both ICL and QLoRA fine-tuning improved LLMs’ performance over zero-shot, and fine-tuning performance was consistently above ICL across all automatic metrics (**Figure 3**). Additionally, we observed substantial limitations of ICL for echo reporting, particularly due to the longer context lengths typical of echo reports. The behavior of integrating results from multiple examples also presents challenges, as discussed below.

Compared to CXR reports, echocardiography reports come with a relatively longer context, which directly affects the available choices of LLMs. In contrast to a prior study that used 32 or more examples(13), we were only able to test up to 4 examples for ICL, therefore not able to assess the models’ behavior with more examples. However, across all metrics, LLMs’ had gradually down-trending performance when more examples were provided (**Figure 3**), which is consistent with prior studies(13,35). It is also important to consider that the computation time and resources required in ICL can increase with the number of examples used(35). Additionally, we note that LLMs can integrate information from the example ICL cases, which compromises the report quality (**Supplemental Table 3**). While this behavior was not reported in other studies(13,35), we believe it could be a relatively common condition, that is easier to identify with numerical values (in echo) compared to narrative statements (in CXR). Therefore, even in scenarios where ICL can outperform fine-tuning(13), fine-tuning may be preferable.

The EchoGen study previously demonstrated that Bidirectional Auto-Regressive Transformers (BART) was superior to other rule-based approaches for summarizing echocardiogram reports, with BART achieving ROUGE-based scores between 0.65 and 0.73, however, human summaries were preferred by the majority of the time over those generated by the BART model(26). Although EchoGPT didn’t match the scores in ROUGE-L, it compared favorably to human experts in qualitative assessments. Additionally, in our study, we observed that T5 (as the representative seq2seq model since EchoGen is not publicly available) generated summaries that were overly brief, so important clinical information was missed (**Supplemental Table 2**). We assume that similar behavior could have occurred with BART, leading to its unfavorable ratings by physicians. However, direct comparisons were not feasible as neither the EchoGen model nor qualitative examples generated by it were available for review.

### Evaluation of Echo Report Summarization

Automatic evaluation of LLM in clinical text summarization tasks is an emerging area, and there is no gold standard metric that can evaluate all aspects of a report(13,26). Our study reinforces this conclusion. We noted that MedAlpaca and Zephyr can generate medical-professionally-sounding content that frequently includes hallucinated information. These differences were mainly reflected by the factual correctness metric RadGraph F1 (**Table 4**).

The practice style at each institution could greatly affect the quantitative performance of a model(18). In the AZ validation set, we observed a 5-10% drop in performance of both fine-tuned Llama-2 and Zephyr (**Supplemental Table 4**). This is likely secondary to the differences in practice style: the AZ validation set, despite having a similar Finding section length, contained an average of 9.5 additional tokens in the Final Impression section (**Table 3**). A more significant drop in performance was observed in reports from the MIMIC-EchoNotes datasets (RadGraph F1 from 47.7 to 25.1; **Supplemental Table 4**), which was anticipated and within a reasonable range(50). According to our observations and the input of expert reviewers, the key factors leading to the performance drop were:

1. The distinct report structure (Findings/Impressions versus Findings/Conclusions/Impressions).
2. The use of reporting languages (templated statements at Mayo versus the free-text style of the MIMIC-EchoNotes dataset).

Specifically, the combination of the Conclusion and Impression sections makes the section almost as long as the Findings (184.3 ± 50.0 vs. 163.2 ± 40.4; **Table 3**), and even longer in some cases. Additionally, the physician’s interpretation often contains information beyond the Findings, or variations of the original sentences, that the model was not able to summarize. Moreover, the sentence templates at Mayo include measurement numbers in relevant statements (e.g., ejection fraction, right ventricular systolic pressure, left atrial size index), while MIMIC-EchoNotes did not (**Table 6**). These factors contributed to the overall less favored completeness, correctness, and clinical utility of EchoGPT summaries on the MIMIC-EchoNotes dataset (**Supplemental Figure 3**). It is important to note the low agreement on correctness and clinical utility metrics, which also implies the challenge on comparing reports with distinct styles (**Supplemental Table 5**). The difference across institutions, as listed above, could limit the direct generalization of a fine-tuned LLM for report summarization. However, while not comprehensively tested in the current study, we noted that adjusting the prompt to fit the reporting style could lead to better summaries without further fine-tuning(50,51).

Regarding the correlations between automatic metrics and human expert preference, our results were similar to the prior studies, showing that most of the metrics were not strongly correlated(13). Notably, the highest correlation was observed between the RadGraph F1 scores and the 4C metrics, particularly in terms of clinical utility (r=0.42) (**Figure 5C**). This suggests that the quality of echo reports judged by cardiologists may not be well captured in automated metrics that do not capture notions of factual correctness. While stronger correlations were observed in the MIMIC-EchoNotes examples (**Supplemental Figure 3**), we believe it was reflecting the strong preference secondary to the distinct reporting style.

As a specific subtype of clinical text, echocardiography reports contain unique terminology, including precise measurements. Clinically, 25% and 55% LV ejection fraction values indicate a significant difference, however, our study demonstrates that this distinction is difficult to capture with current automatic metrics (**Table 5**). While this aspect of reporting can be easily captured in qualitative analyses, such analyses are expensive to conduct at scale because of the limited availability of in-domain experts. A dedicated metric for echocardiography diagnostic quality evaluation, with emphasis on measurement accuracy, is still needed to address this knowledge gap.

### Application of EchoGPT

Our study shows the feasibility of introducing LLMs into echocardiography practice. Through the QLoRA fine-tuning process(38), the EchoGPT model was able to learn clinically relevant knowledge to summarize echo report findings at a quality level comparable to echocardiography-trained cardiologists (**Figure 6A**).

We envision that EchoGPT could be used as a reporting interface or a co-pilot that could generate echo reports with various inputs(52). EchoGPT still inherits the limitations of LLMs, including hallucination(13,18,53). Although the fine-tuning process can potentially reduce hallucinations, additional efforts such as optimization for factual correctness(46) or paired with a retrieval augmented generation system(54) are still required to minimize hallucinations before clinical implementation.

## Conclusion

Our study successfully built EchoGPT through QLoRA fine-tuning of open-source LLMs and demonstrated that the model is capable of generating echocardiography reports on par with cardiologists, marking an advancement in integrating LLMs into current echo practice. Our analysis also highlights the challenges of the current LLM evaluation process in echo reporting, particularly dedicated automatic metrics and scalable qualitative evaluation approaches, which necessitate further studies.

## Limitations

This study is limited by its retrospective nature and a predominantly white population served by the healthcare system. However, we were able to demonstrate that the algorithm is not biased by sex and race. Our echocardiography reports are based on standardized statements, with an option to add free text. While the lexical variance was high, the corpus could differ from reports composed entirely of free text contents. Due to patient privacy regulations, this work did not assess the performance of GPT-3.5 and GPT-4.

However, we compared the performance of state-of-the-art open-source LLMs, which provided important insights for model selection when data privacy is a critical consideration. Instead of full fine-tuning, QLoRA was used as the fine-tuning approach, however, it has been demonstrated as an effective approach as full fine-tuning is often not feasible for LLMs. Last but not the least, although QLoRA fine-tuning demonstrated improvements in echo report summarization tasks, our current approach does not include optimization for factual correctness and human expert preference.

## Data Availability

The data that support the findings of this study are not openly available due to reasons of sensitivity and patient privacy. Data are located in controlled access data storage at the Mayo Clinic. The MIMIC-EchoNotes (ECHO-NOTE2NUM) data is publicly available at https://doi.org/10.13026/xhrz-ht59

## Code Availability

We released a checkpoint of the fine-tuned Llama-2 model, along with the QLoRA fine-tuning, inference, and statistical analysis code. The code and checkpoint are available on GitHub: https://github.com/chiehjuchao/EchoGPT.git

---

**Supplemental Figure 1.**
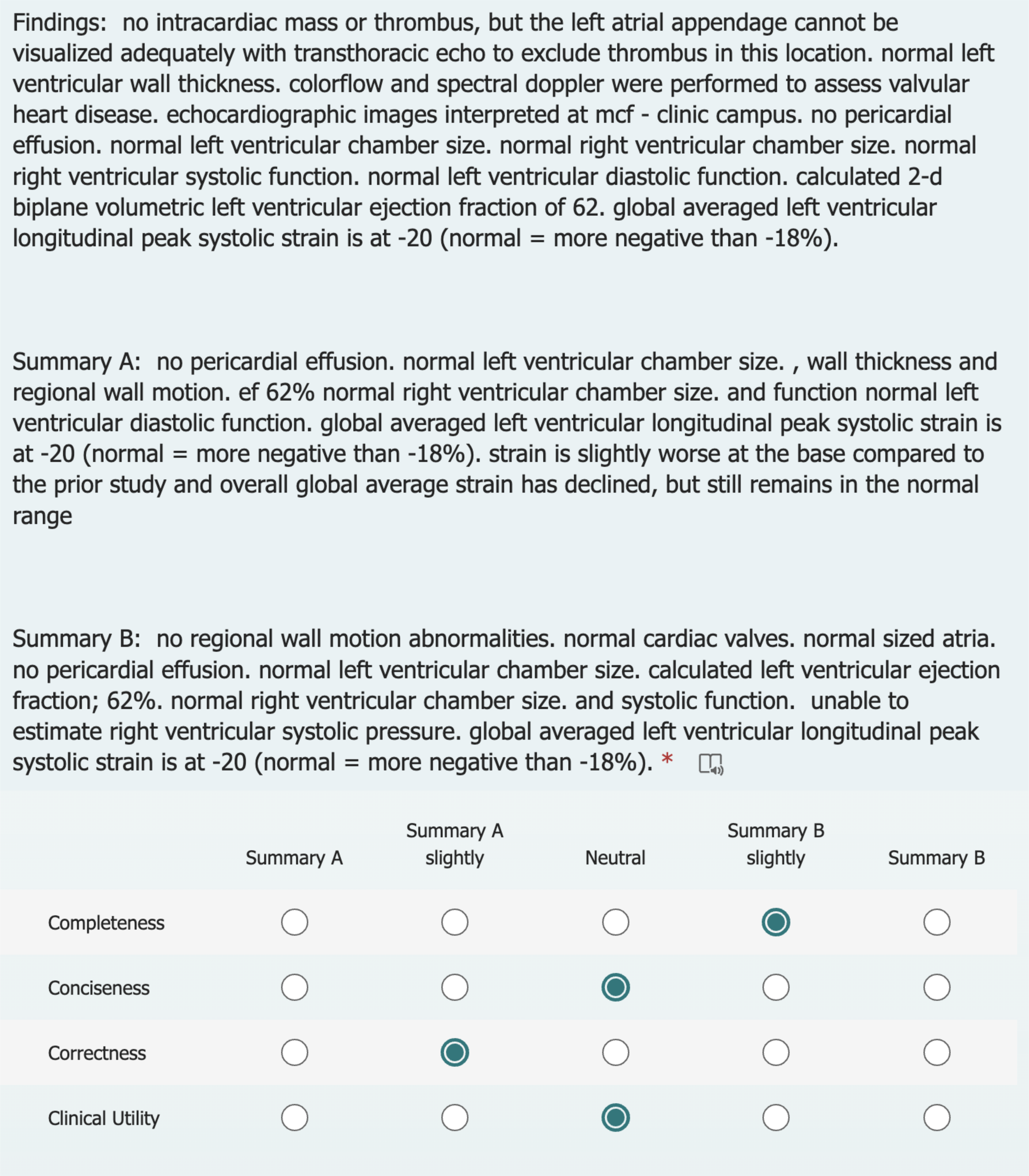
Echocardiography expert review questionnaire. Expert readers were asked to rate summaries A and B concerning the 4C metrics without knowing it’s a human-or LLM-generated summary.

**Supplemental Figure 2.**
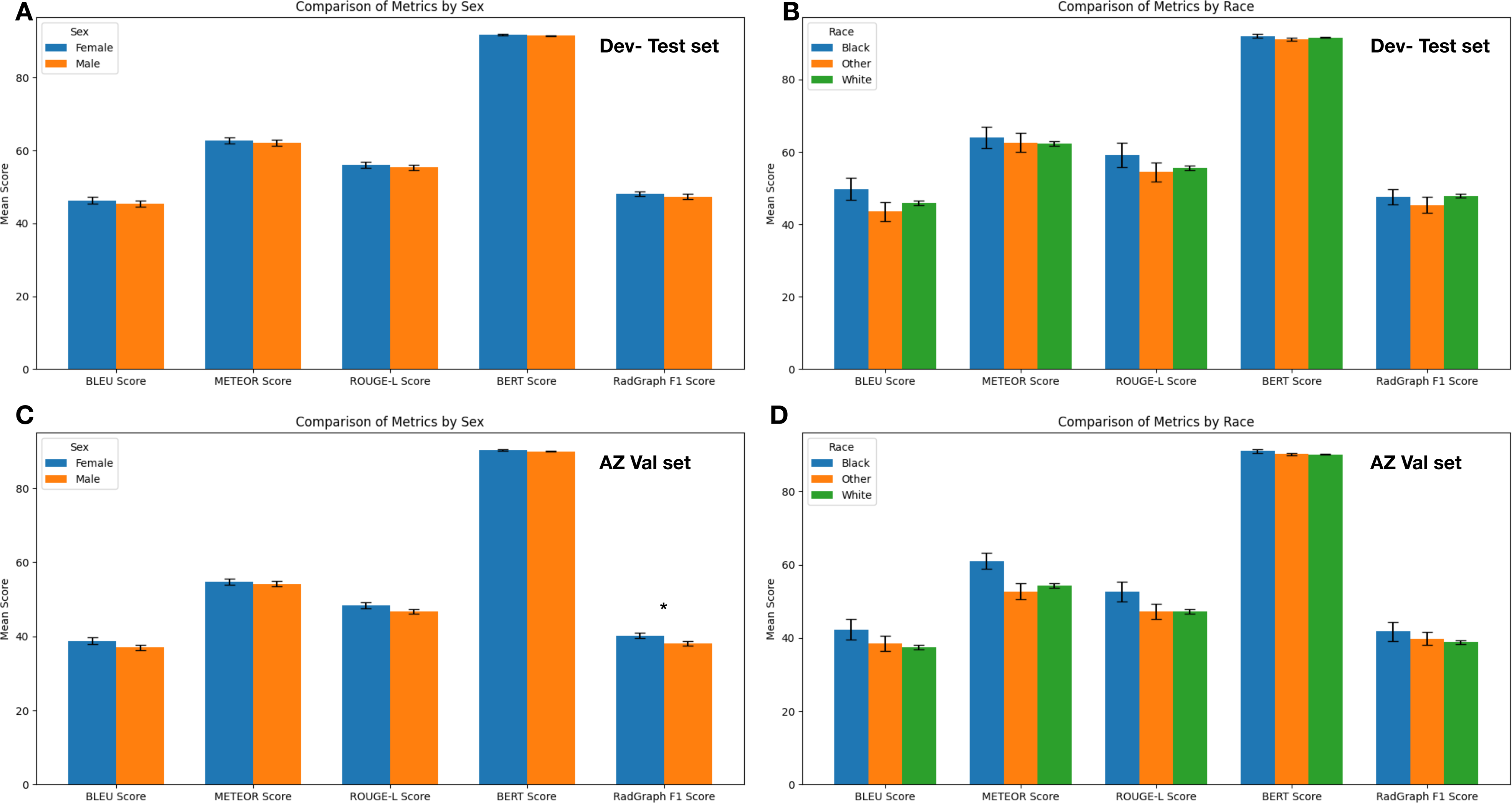
This bar chart displays model performance across five automatic metrics, considering sex (male vs. female) and race (white, black, and other) variables. Panels A and B compare metrics by sex and race variables in the test set, while Panels C and D perform the same comparisons in the AZ validation set. There were no significant biases detected for sex or race, except for slightly better RadGraph F1 performance in female patients within the AZ validation set (male vs. female: 0.38 ± 0.14 vs. 0.40 ± 0.15, p=0.04). Scores were presented as mean values ± standard error bars. Demographic information was not available for the same analysis in the MIMIC-EchoNotes dataset.

**Supplemental Figure 3.**
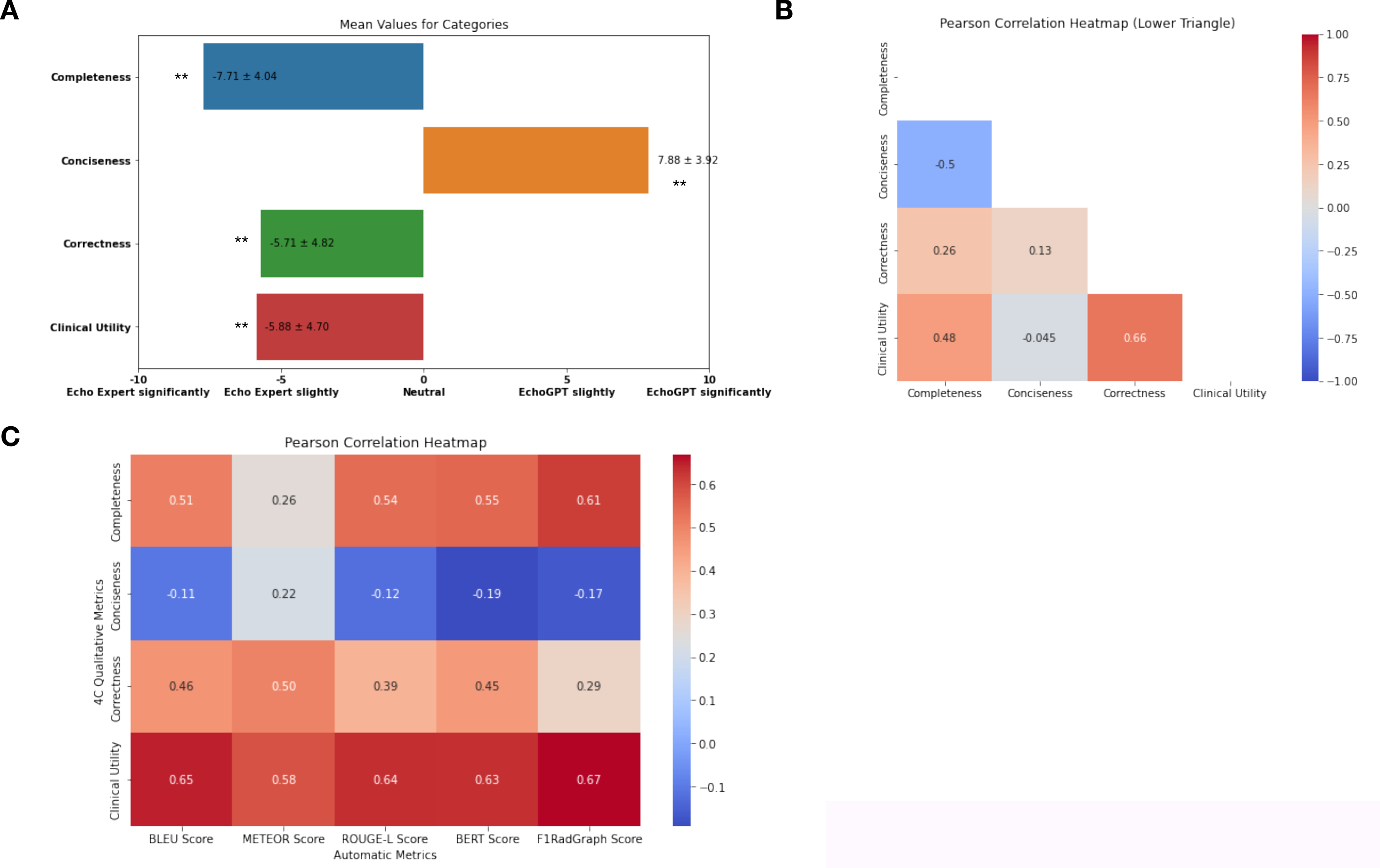
Human expert qualitative evaluation results on the MIMIC-EchoNotes dataset. **Panel A**. In the 4 categories, the original reports (combined Conclusions and Impression sections) were preferred over EchoGPT in completeness, correctness, and clinical utility (p< 0.001); while EchoGPT was superior in conciseness (p<0.001). **Panel B.** Interdependence of the 4C metrics, especially the correlations between clinical utility, completeness, and correctness (Pearson’s r= 0.48 and 0.66, respectively). **Panel C.** Correlations between automatic metrics and the 4C metrics. Across all automatic metrics, RadGraph F1 still had the strongest correlations with the 4C metrics, although other metrics also had stronger correlations. Preference ratings were expressed as mean ± standard deviation, **indicates p<0.001.

**Supplemental Table 1.** Hyperparameter Search Results.

|  | Temperature (RP fixed at 1.1) |  |  | Repetition Penalty (Temp fixed at 0.1) |  |
| --- | --- | --- | --- | --- | --- |
|  | 0.1 | 0.5 | 0.9 | 1.2 | 1.3 |
| **LLaMA-2** |  |  |  |  |  |
| BLEU | 8.0 ± 5.4 | 8.8 ± 6.7 | 8.0 ± 6.1 | 5.5 ± 4.6 | 1.1 ± 1.8 |
| METEOR | 23.3 ± 8.2 | 24.1 ± 8.2 | 23.0 ± 8.1 | 20.1 ± 6.9 | 12.9 ± 5.7 |
| ROUGE-L | 22.3 ± 7.4 | 23.2 ± 7.8 | 23.3 ± 9.3 | 18.7 ± 7.0 | 11.6 ± 5.3 |
| BERT Score | 85.7 ± 1.5 | 86.0 ± 1.4 | 85.8 ± 1.6 | 84.7 ± 2.0 | 82.4 ± 1.6 |
| RadGraph F1 | 26.2 ± 9.9 | 25.9 ± 10.7 | 26.2 ± 12.4 | 20.4 ± 9.9 | 10.3 ± 7.0 |
| **MedAlpaca** |  |  |  |  |  |
| BLEU | 0.9 ± 2.8 | 0.5 ± 1.6 | 0.7 ± 2.0 | 0.1 ± 0.8 | 0.0 ± 0.0 |
| METEOR | 12.3 ± 7.4 | 13.9 ± 8.9 | 13.1 ± 9.4 | 9.2 ± 6.1 | 5.6 ± 3.2 |
| ROUGE-L | 9.9 ± 7.3 | 10.4 ± 7.9 | 9.4 ± 7.5 | 6.7 ± 4.9 | 3.9 ± 2.7 |
| BERT Score | 81.3 ± 2.7 | 81.6 ± 2.8 | 81.4 ± 3.1 | 80.4 ± 2.2 | 79.4 ± 1.6 |
| RadGraph F1 | 7.1 ± 8.2 | 7.4 ± 7.1 | 7.9 ± 8.3 | 3.2 ± 3.9 | 1.0 ± 2.1 |
| **Zephyr** |  |  |  |  |  |
| BLEU | 4.4 ± 6.3 | 4.0 ± 4.5 | 4.4 ± 5.0 | 1.3 ± 2.7 | 0.1 ± 0.6 |
| METEOR | 23.5 ± 8.3 | 22.8 ± 7.6 | 22.6 ± 9.1 | 18.1 ± 6.6 | 12.8 ± 5.8 |
| ROUGE-L | 20.2 ± 7.8 | 19.6 ± 7.4 | 19.0 ± 7.6 | 14.6 ± 5.5 | 10.6 ± 5.1 |
| BERT Score | 83.6 ± 2.8 | 83.9 ± 2.5 | 83.5 ± 2.9 | 82.8 ± 2.0 | 81.5 ± 1.7 |
| RadGraph F1 | 16.5 ± 9.6 | 16.4 ± 9.8 | 14.8 ± 10.8 | 7.9 ± 7.0 | 3.3 ± 4.7 |
| **Flan-T5** |  |  |  |  |  |
| BLEU | 9.6 ± 9.3 | 10.3 ± 10.2 | 9.8 ± 8.7 | 9.3 ± 8.2 | 9.9 ± 9.3 |
| METEOR | 37.0 ± 14.8 | 37.9 ± 15.3 | 37.5 ± 15.0 | 37.8 ± 15.0 | 37.7 ± 14.0 |
| ROUGE-L | 22.8 ± 10.2 | 23.2 ± 10.4 | 22.7 ± 9.6 | 22.6 ± 9.8 | 22.8 ± 10.0 |
| BERT Score | 85.8 ± 2.2 | 86.1 ± 2.2 | 85.9 ± 2.3 | 86.0 ± 2.3 | 85.9 ± 2.1 |
| RadGraph F1 | 18.2 ± 10.3 | 19.5 ± 10.9 | 18.3 ± 10.0 | 17.9 ± 10.4 | 18.5 ± 9.7 |
| Temp: temperature, Repetition Penalty: RP, BLEU: Bilingual Evaluation Understudy, METEOR: Metric for Evaluation of Translation with Explicit ORDERing, ROUGE-L: Recall-Oriented Understudy for Gisting Evaluation - Longest Common Subsequence, BERT: Bidirectional Encoder Representations from Transformers. |  |  |  |  |  |

**Supplemental Table 2.** Representative Example of Zero-shot Results.

| Model | Ground Truth | LLaMA-2 | Flan-T5 | Zephyr | MedAlpaca |
| --- | --- | --- | --- | --- | --- |
| **Representative Example** | Global hypokinesia with superimposed regional variation (please see wall motion graphics). estimated right ventricular systolic pressure 32 (systolic blood pressure 120). no pericardial effusion. no significant valvular heart disease. moderate-severely enlarged left ventricular chamber size; calculated ejection fraction 25%. normal right ventricular chamber size. with borderline decrease in systolic function. with normal inspiratory collapse (>50%) inferior vena cava size normal. indeterminate left ventricular diastolic function." | The patient presents with global hypokinesia and regional variation in left ventricular wall motion. There is no evidence of intracardiac mass or thrombus, but the left atrial appendage cannot be adequately visualized for thrombus exclusion. The estimated right ventricular systolic pressure is 32 mmHg, and there is an anterior epicardial fat layer. The pulmonary valve is partially visualized, and the abdominal aorta has an incomplete Doppler flow pattern. The left ventricular wall thickness is normal, and there is no atrial level shunt. However, the left atrium volume index is slightly elevated, and there is mild mitral valve regurgitation and trivial pulmonary valve regurgitation. Additionally, the patient has moderate-severe left ventricular chamber enlargement, normal right ventricular chamber size, and normal aortic valve function. Overall, the patient's cardiac function appears compromised, likely due to chronic heart failure. Further evaluation and management are warranted. | Calculated 2-d linear left ventricular ejection fraction 25. global hypokinesia with superimposed regional variation. No intracardiac mass or thrombus. | LVEF: 25% with superimposed regional variation (hypokinesia) - No intracardiac mass, but LAA cannot be ruled out for thrombus (transthoracic only) - Mild MR; trivial TR and PVR - Anterior epicardial fat; pulmonary valve not well visualized - Normal RV systolic pressures, AV, and RA sizes - Moderate-severe chamber enlargement; borderline RV syst. | Summary: This is a good study from which I can conclude that the patient has mild MR and TR. His LVESD is slightly above normal and his RVSV is slightly decreased. His PVR is also low. He does have a thrombus in the left atrial appendage, although it may not be noticeable on this particular study. His pulmonary artery velocity is normal but the abdominal aorta could not be measured adequately. So more information is needed for a full diagnosis. There is some fat in the epicardium, which is also nonspecific and might be seen in many patients. There is evidence of LV strain but again, more information is needed. The LA size is normal as is the RV size, and the IVC was normal. The aorta was normal in size. In general, there are no significant abnormalities and no cause for concern at this time. |
| Comments | -- | Summary of the findings with transitioning words that did not alter the original meaning. | Concise summary but omitted clinical important information. | Concise bullet point summary with false information. | Medical Professional-sounding, but hallucinated information. |

**Supplemental Table 3.** Representative Example: In-context Learning Integrated Information from Multiple Examples.

|  | Sentence | Model Summary |
| --- | --- | --- |
| Case | Calculated 2-d linear left ventricular ejection fraction 57. | "Calculated 2-d linear left ventricular ejection fraction 57, 61, 75 (3 reports)" |
| ICL Examples | Calculated 2-d linear left ventricular ejection fraction 56. |  |
|  | Calculated 2-d linear left ventricular ejection fraction 75 |  |
|  | Calculated 2-d linear left ventricular ejection fraction 69 |  |
|  | Calculated 2-d linear left ventricular ejection fraction 61 |  |

**Supplemental Table 4.** Performance of Fine-tuned LLMs on the AZ Validation and MIMIC-EchoNote Datasets.

| Dataset | Mayo AZ validation |  |  | MIMIC-EchoNote |  |  |
| --- | --- | --- | --- | --- | --- | --- |
| Model | LLaMA-2 | Zephyr |  | LLaMA-2 | Zephyr |  |
|  | QLoRA | QLoRA | P-value* | QLoRA | QLoRA | P-value* |
| BLEU | 37.8 ± 17.5 | 17.6 ± 13.8 | <0.0001 | 9.6 ± 10.6 | 3.6 ± 4.5 | <0.0001 |
| METEOR | 54.4 ± 17.3 | 32.9 ± 14.9 | <0.0001 | 40.2 ± 13.3 | 23.3 ± 10.1 | <0.0001 |
| ROUGE-L | 47.4 ± 16.9 | 29.1 ± 13.7 | <0.0001 | 25.1 ± 12.8 | 14.6 ± 7.2 | <0.0001 |
| BERT Score | 90.1 ± 2.9 | 86.6 ± 3.8 | <0.0001 | 85.4 ± 2.0 | 80.2 ± 15.2 | <0.0001 |
| RadGraph F1 | 39.0 ± 14.5 | 25.4 ± 12.2 | <0.0001 | 25.1 ± 13.9 | 15.4 ± 9.8 | <0.0001 |
| *Compared performance of QLoRA Llama-2 and Zephyr |  |  |  |  |  |  |

**Supplemental Table 5.** Agreement of the Ratings Between Echo-Expert Readers.

| Metric | Fleiss' Kappa between 4 raters |  |
| --- | --- | --- |
|  | Mayo | MIMIC-Echo |
| Completeness | 0.49 | 0.49 |
| Conciseness | 0.22 | 0.60 |
| Correctness | 0.17 | 0.12 |
| Clinical Utility | 0.34 | 0.14 |

**Central Illustration.**
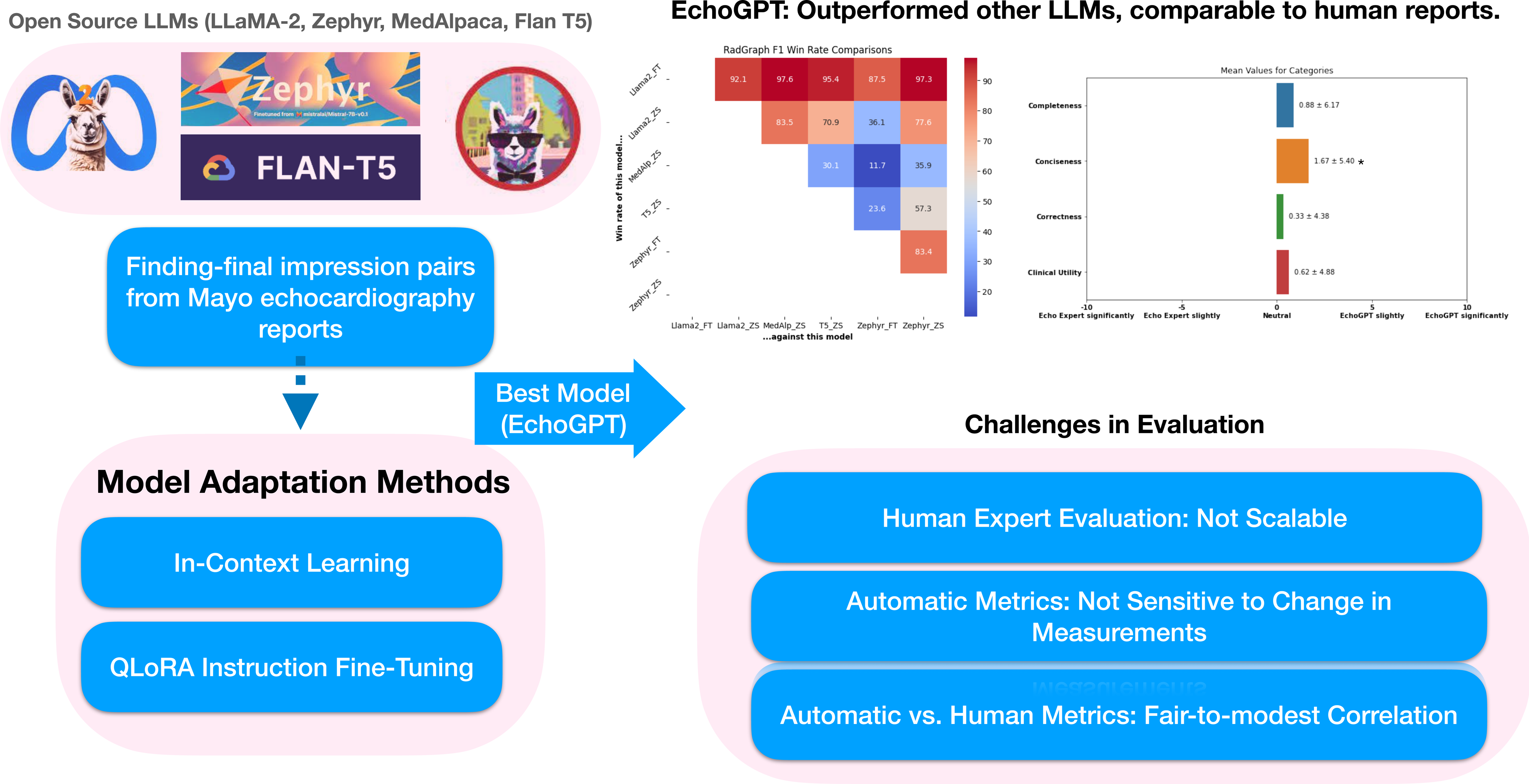
Open-source LLMs (Llama-2, MedAlpaca, Zephyr, and Flan-T5) were evaluated using In-Context Learning (ICL) and Quantized Low-Rank Adaptation (QLoRA) fine-tuning for summarizing echocardiography reports from “Findings” to “Impressions.” EchoGPT, a QLoRA fine-tuned Llama-2 model, surpassed other LLMs in multiple automatic metrics and produced reports on par with those of cardiologists in qualitative reviews. However, challenges in evaluating generated reports included limited scalability of expert reviews, modest correlations between automatic and human metrics, and the automatic metrics’ insensitivity to changes in measurements.

## Abbreviations

4C metrics: qualitative review metrics including completeness, correctness, conciseness, and clinical utility.
AI: Artificial Intelligence
DL: Deep Learning
Echo: Echocardiography
FT: Fine-Tuning
GPT: Generative Pre-trained Transformer
ICL: In-Context Learning
NLP: Natural Language Processing
Seq2seq: Sequence-to-sequence
TTE: Transthoracic Echocardiography
TEE: Transesophageal Echocardiography
LLM: Large Language Model
NLP: Natural Language Processing
QLoRA: Quantized Low-Rank Adaption

